# A Patient-Specific CFD Study of Carotid Webs: Hemodynamic Analysis and the Role of Blood Viscosity

**DOI:** 10.64898/2026.03.18.26348736

**Authors:** Xuning Zhao, Farhan Khan, Skylar Lewis, Mauro Rodriguez

## Abstract

**Background:** Carotid webs (CaWs) are shelf-like protrusions in carotid bifurcation recognized as a potential cause of ischemic stroke. However, their impact on wall-based hemodynamic metrics (TAWSS, OSI, RRT) in distinguishing from normal bifurcations remains unclear.

**Methods:** Carotid geometries were reconstructed from CT angiography in patients with CaWs, classified as symptomatic (with ischemic stroke) or asymptomatic (incidentally detected), and controls with normal bifurcations. Influence of three blood viscosity models (Newtonian, Carreau–Yasuda, Casson) was evaluated. Metrics were quantified using a Gaussian-weighted spatial averaging method and compared between groups.

**Results:** CFD simulations were performed in 22 CaW cases (16 symptomatic, 6 asymptomatic) and 6 normal bifurcations. Simulations predicted recirculation corresponding to delayed contrast clearance on DSA. Viscosity models had minimal influence on flow patterns (<2% differences). CaWs showed greater inter-patient variability than normal bifurcations, but overlap remained (e.g., TAWSS 3.39 (2.72–8.96) vs 4.18 (3.09–4.56) Pa, *p* = 0.858). Symptomatic CaWs showed lower TAWSS and higher OSI and RRT than asymptomatic CaWs (TAWSS 3.39 vs 6.63 Pa), although didn’t reach statistical significance (*p* > 0.25).

**Conclusion:** Symptomatic CaWs show lower shear stress and stronger oscillations than asymptomatic CaWs. However, wall-based hemodynamic metrics alone may not distinguish CaWs from normal carotid geometries.

## 1. Introduction

Carotid webs (CaWs) are thin, fibrous, shelf-like intimal protrusions that typically arise from the posterior wall of the internal carotid artery (ICA)^1–3^. Recently, CaWs have emerged as an under-recognized cause of ischemic stroke, particularly in younger patients without traditional vascular risk factors and in the absence of high-grade stenosis^3–5^. Recent studies report that CaWs may account for up to 30% of cryptogenic strokes in patients younger than 60 years^1,4,6^, and a stroke recurrence risk as high as 20% within two years despite medical management^7–9^. Rigorously characterizing the hemodynamics will inform how CaWs contribute to stroke risk, find patients at elevated risk, and inform individualized treatment strategies.

Local hemodynamics are increasingly recognized as a key factor linking CaW to ischemic stroke^3–5^. The shelf-like projection disrupts flow in the carotid artery, generating regions of separation, recirculation, and stasis distal to the lesion. These abnormal flow environments promote thrombus formation and increase the likelihood of embolization^2^. For example, catheter angiography or digital subtraction angiography (DSA) shows contrast stagnation or delayed clearance downstream of CaW, consistent with recirculating flow^3,4,10,11^. However, DSA is invasive, projection-based imaging modality providing qualitative indications of hemodynamics. El Sayed et al. (2023)^12^ used three-dimensional, three-directional, ECG-gated, time-resolved, phase-contrast magnetic resonance imaging (4D Flow MRI) to measure time-resolved velocity fields in patients with CaW. The imaging revealed reduced near-wall velocities, oscillatory flow, and recirculation zones. However, 4D Flow MRI has low spatial and temporal resolution, limiting its ability to capture small-scale features (e.g., near-wall velocities and subtle geometric variation around the web) necessary to characterize CaW-related flow disturbances.

Computational fluid dynamics (CFD) enables the detailed study of CaW hemodynamics by resolving full spatial transient flow fields and enabling the computation of detailed hemodynamic quantities of interest. Since CaWs have only recently gained clinical recognition, the number of CFD studies is small^13–19^. The studies consistently suggest that CaWs generate disturbed flow with separation, recirculation, and low wall shear stress downstream of the lesion. However, direct validation of these predictions against clinical or experimental measurements remains scarce. These studies employed artificially constructed or patient-specific geometries for the carotid artery with CaW. Studies using artificially constructed geometries are useful for isolating the effects of specific web features but do not capture the anatomical heterogeneity seen in real patients. Patient-specific studies reflect the real individual anatomy^16–19^, but they are typically limited to small sample sizes and lack consistent strategies for cross-patient comparison. The treatment of blood viscosity also varies across existing studies, using either constant Newtonian viscosity^17,18^ or Carreau-Yasuda^13–16^. Blood exhibits shear-dependent viscosity at low shear conditions (i.e., <10^2^ 1/s) due to red blood cell aggregation and deformation, with apparent viscosity decreasing as shear rate increases^20–22^. Since CaWs introduce disturbed flow conditions and low shear rate downstream of the lesion^17^, the choice of viscosity model may influence hemodynamic predictions. However, it remains unclear how different viscosity models influence key hemodynamic features in CaWs.

To characterize the hemodynamics of CaWs, prior studies as well as broader vascular research^23–26^, have focused on wall shear stress (WSS), its temporal average over the cardiac cycle (time-averaged WSS, TAWSS), oscillatory shear index (OSI), and the relative residence time (RRT) of blood. WSS and TAWSS indicate the overall magnitude of frictional forces induced by the blood flow acting on the vessel wall. OSI quantifies the degree of directional change in wall shear over the cardiac cycle, and RRT describes the persistence of blood elements near the wall^27,28^. Low shear, high OSI, and prolonged near-wall residence are commonly regarded as markers of disturbed flow environments that promote thrombus formation^29–31^. These metrics are widely applied as surrogates of pro-thrombotic environments^28,32^, but it is still unknown if such features are sufficiently effective to differentiate CaW from normal bifurcations. Two strategies have been used when comparing patient-specific geometries with heterogeneous anatomical features. On the one hand, local extrema measures^16^ quantify maximum (or minimum) values of hemodynamic indices (e.g., peak OSI, minimum TAWSS). The maxima are inherently unstable, driven by single points, sensitive to mesh/temporal resolution, and prone to outliers, so they may not reflect the overall hemodynamic patterns. On the other hand, region-of-interest (ROI) averaging^17^ metrics are averaged within a predefined vascular segment (e.g., a fixed-length window). These metrics offer a broader characterization but depend on how the ROI is chosen and may dilute highly localized disturbances, complicating cross-patient comparability.

The objective of this study is to investigate the hemodynamic characteristics of carotid webs using patient-specific computational fluid dynamics. We introduce a Gaussian-weighted spatial averaging approach to quantify wall-based hemodynamic metrics and enable comparison across different geometries. Using this framework, we evaluate the influence of different blood viscosity models and compare hemodynamic metrics between carotid webs and normal carotid bifurcations, as well as between carotid webs associated with ischemic stroke (symptomatic CaWs) and incidentally detected carotid webs (asymptomatic CaWs).

## 2. Material and Methods

### 2.1 Patient cohort and imaging data

The study was approved by the Institutional Review Board (IRB), and the imaging data were de-identified prior to analysis. Given the retrospective nature of the study, the requirement for informed consent was waived. The data that supports the findings of this study can be obtained from the corresponding author upon reasonable request.

This retrospective observational study included patients with carotid artery webs (CaWs), both symptomatic and asymptomatic, along with age-matched controls without evidence of atherosclerosis or ipsilateral ischemic stroke admitted at our institution. CaWs were defined as thin, shelf-like linear projections arising from the posterior wall of the internal carotid artery (ICA), visualized on both axial and sagittal imaging planes in the absence of atherosclerosis. The diagnosis of CaW was established by a board-certified neuroradiologist, vascular neurologists and neurointerventionalists, with webs defined as thin, shelf-like projections from the posterior wall of the proximal internal carotid artery, visualized on both axial source images and confirmed on multiplanar reconstructions. We excluded patients with superimposed thrombus on CaW. CT angiography (CTA) of the neck was used to identify CaWs and controls, performed according to institutional protocol (see Supplemental Materials). Continuous variables are reported as means or medians, and categorical variables as proportions. Continuous variables were compared using Mann Whitney U test, and categorical variables were analyzed using the Kruskal–Wallis test and the Wilcoxon rank-sum test.

### 2.2 Pre-processing

Segmentation of the vascular lumen was performed in the open-source 3D Slicer (version 5.6.2) using semi-automated methods with manual refinement. The segmented domain included the common carotid artery, carotid bifurcation, external carotid artery, and proximal internal carotid artery. The constructed lumen surfaces were exported in STL format and cropped to include 30-40 mm of vessel length both upstream and downstream of the region of interest, ensuring sufficient flow development for computational modeling. The geometries were verified by the neuroradiologist before meshing. An example of the segmentation and resulting reconstructed geometry is shown in Figure 1a and 1b.

**Figure 1.**
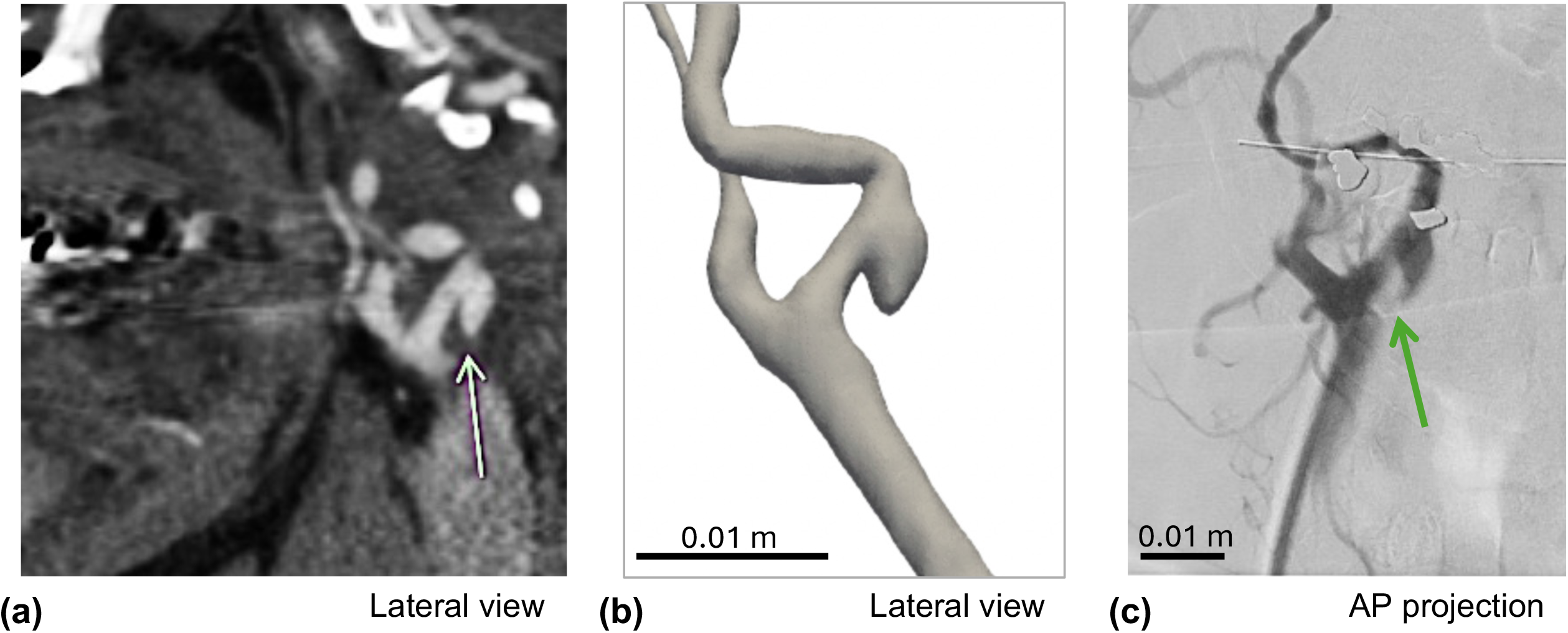
(a) Axial CTA image showing a carotid web as a thin, shelf-like protrusion arising from the posterior wall of the proximal internal carotid artery (arrow). (b) Reconstructed 3D lumen geometry derived from CTA segmentation. (c) DSA of the same carotid bifurcation in an anteroposterior projection, confirming the presence and location of the CaW (arrow).

Catheter-based digital subtraction angiography (DSA) was used in a subset of patients as part of routine clinical care. DSA was performed using standard intra-arterial contrast injection and biplanar acquisition protocols. For patients with a CaW, DSA served both as a reference modality for confirming the presence of the web (Figure 1c) and qualitative validation of simulated flow features (see Results).

### 2.3 Computational Fluid Dynamics Simulation

The STL geometry files were imported into OpenFOAM (Version: v2206) for geometry meshing and transient flow simulations. Volumetric meshes of unstructured tetrahedral elements were generated using the *snappyHexMesh* utility. A mesh-independence resolution study was performed using the WSS as the convergence criterion. The time-spatially averaged WSS converged at mesh resolution of approximately 4 million elements (see Supplementary Figure S1a). Subsequent simulations use mesh resolutions beyond 4 million elements.

Blood flow was modeled as a viscous fluid governed by the incompressible Navier-Strokes equations, with a density of 1060 kg/m^3^. The governing equations were solved using the *pimpleFOAM* transient solver in OpenFOAM. Vessel walls were assumed to be rigid with a no-slip boundary condition. At the inlet of the common carotid artery, a pulsatile volumetric flow waveform was prescribed based on published flow data in Lee et al. (2008)^33^. The waveform was rescaled according to the patient-specific heart rate, and the resulting inflow condition corresponding to a cardiac cycle of 85 bpm is shown in Supplementary Figure S1b. The velocity distribution at the inlet was imposed using a plug-like profile. Simulations were run for three cardiac cycles to ensure periodic convergence, and results were analyzed from the final cycle. Traction-free (zero-pressure) boundary condition was applied to both the internal and external carotid artery outlets.

### 2.4 Viscosity models

We considered three different viscosity models: Newtonian, Carreau–Yasuda, and Casson. The Newtonian model assumes a constant, shear-independent viscosity (*μ*_0_ *=* 3.5 × 10^−3^ Pa s). The Carreau-Yasuda model captures the shear thinning behavior of blood at physiological shear rate, 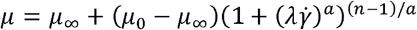, where 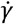 is the shear rate, the relaxation time (*λ* = 3.31 s, parameter *a* = 1, power-law exponent *n* = 0.357, zero-shear-rate viscosity *μ*_0_ 5.6 × 10^−2^ Pa s, and infinite-shear-rate viscosity *μ* = 3.45 × 10^−3^ Pa s. The Casson model incorporates both shear-thinning and yield stress effects, 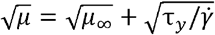, where the yield stress is *τ*_*y*_ = 7.47 × 10^−3^ Pa, and *μ*_∞_ = 3.2 × 10^−3^ Pa s. The predictions of these three viscosity models show good agreement with experimental measurements of blood viscosity over shear rate of 10-1000 s^-1^ reported in multiple studies (see Supplementary Figure S1c).^34–38^

### 2.5 Hemodynamic metrics and cross-patient comparison

Flow fields were post-processed using ParaView (Kitware, Clifton Park, NY, USA) and custom OpenFOAM utilities. Hemodynamic parameters, including time-dependent velocity, WSS, time-averaged WSS (TAWSS), OSI, and RRT were extracted from the final cardiac cycle of each simulation. RRT was calculated using a viscosity-inclusive formulation:

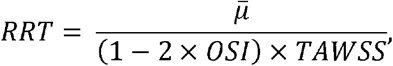

where 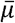 denotes the time-averaged viscosity.

We introduce a weighted spatial averaging formulation for wall-based hemodynamic quantities, i.e., TAWSS, OSI, and RRT. The formulation localizes these quantities with a continuous weighting function centered at the carotid web throat, avoids manual definition of a region of interest, and smoothly down-weighs distant contributions. For a wall-based quantity *q*(*x*), the weighted spatial average value over the entire vessel wall is defined as

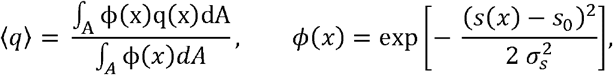

where *A* is the vessel wall surface, *dA* the local surface area element at position *x*, and *ϕ* (*x*) a Gaussian weighting function centered at the CaW throat. *s*(*x*) denotes the projection point of *x* to the vessel centerline, and *s*_0_ the centerline location of the minimum cross-sectional throat area in the CaW (see Figure 2). The parameter *σ*_S_ is the longitudinal extent of the weighted region. We set 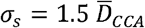, corresponding to a window of approximately 1.5 times the average CCA diameter upstream and downstream of the lesion. For each patient, a single weighted spatially averaged value of each hemodynamic metric was obtained.

**Figure 2.**
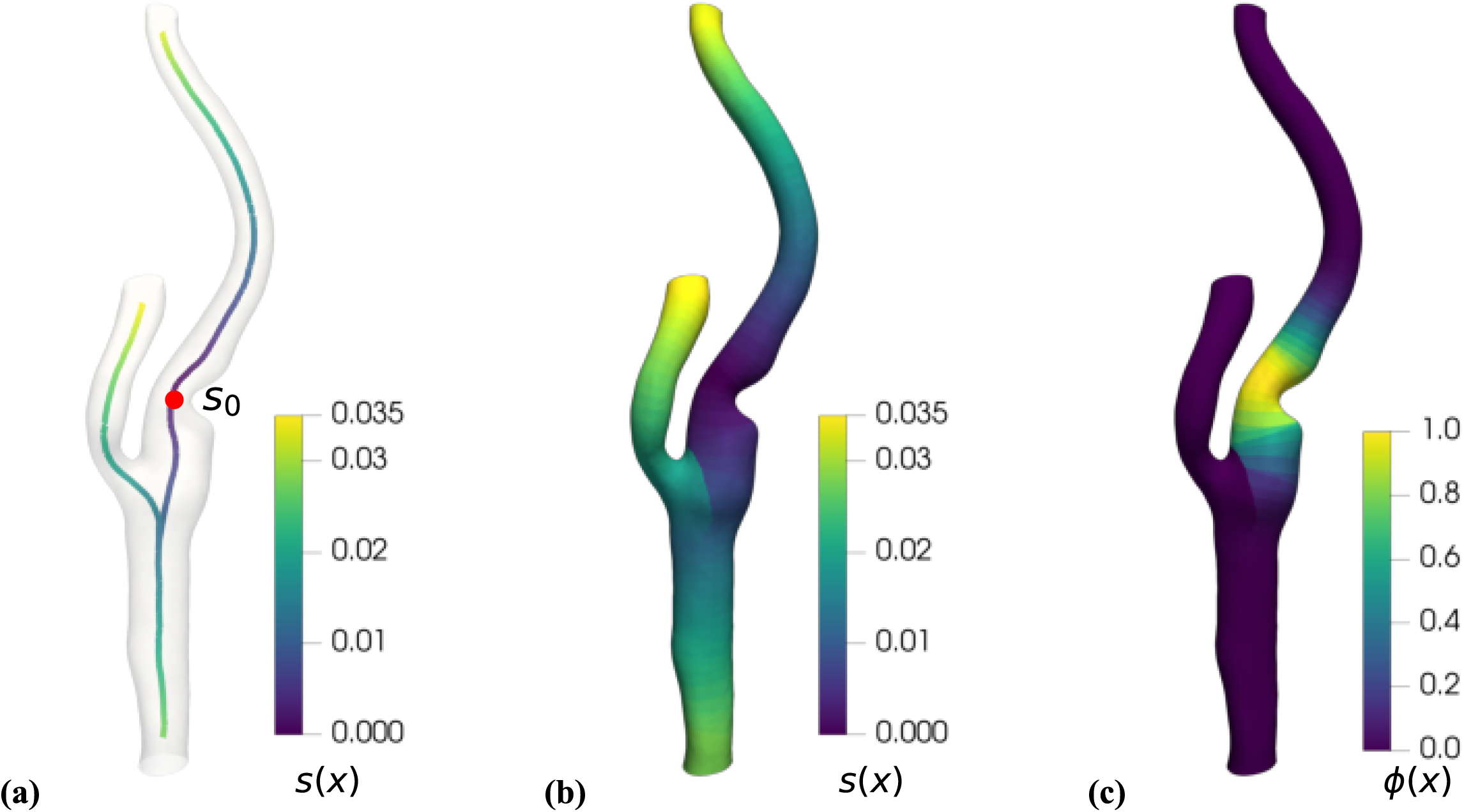
Schematic illustration of the weighted spatial averaging method for cross-patient hemodynamic quantification. (a) Vessel centerline with projection coordinate *s*(*x*) and throat location *s*_0_, defined as the position of minimum cross-sectional area in the CaW region. (b) Projection coordinate *s*(*x*) mapped onto the vessel wall surface. (c) Gaussian weighting function *ϕ* (*x*).

## 3. Results

### 3.1 Patient characteristics

The study included a total of 28 patients: 22 with CaWs, 6 with asymptomatic CaWs, and 6 with normal carotid bifurcations. The median age was 52 years (IQR 48–62) in the CaW group and 54.5 years (IQR 42.0–65.0) in the control group. The majority of patients in the CaW group were female (59%) compared with the control group (33%). The degree of luminal narrowing caused by CaWs ranged from 10% to 40% using NASCET criteria^39^. Demographic and clinical information for both groups is summarized in Table 1.

**Table 1.** Patient characteristics for groups with and without carotid web. Values are reported as median (interquartile range) or number (%).

| Characteristic | Carotid Webs (n = 22) | Normal (n = 6) | p-value |
| --- | --- | --- | --- |
| Age, median (IQR) | 52.0 (48.0, 62.0) | 54.5 (42.0, 65.0) | 0.95 |
| Gender, male | 9 (43%) | 4 (67%) | 0.30 |
| Race, Caucasians | 16 (76%) | 5 (83%) | 0.084 |
| Ethnicity, non-Hispanic | 22 (100%) | 6 (100%) | - |
| Hypertension, yes | 6 (30%) | 2 (33%) | 0.88 |
| Diabetes mellitus, yes | 2 (10%) | 1 (17%) | 0.62 |
| Dyslipidemia, yes | 4 (19%) | 3 (50%) | 0.13 |

### 3.2 Flow fields

Figure 3 shows simulated time-averaged velocity fields for two representative CaW cases and corresponding patient DSA images. In both cases, DSA shows a region of delayed contrast clearance distal to the web (red box in frame a and c), showing reduced through-flow in this segment. The CFD streamlines show a low-velocity region and a downstream recirculation zone at the same axial location (green box in frame b and d). The spatial extent of the simulated low-flow region is comparable to the region of delayed contrast transport seen on DSA.

**Figure 3.**
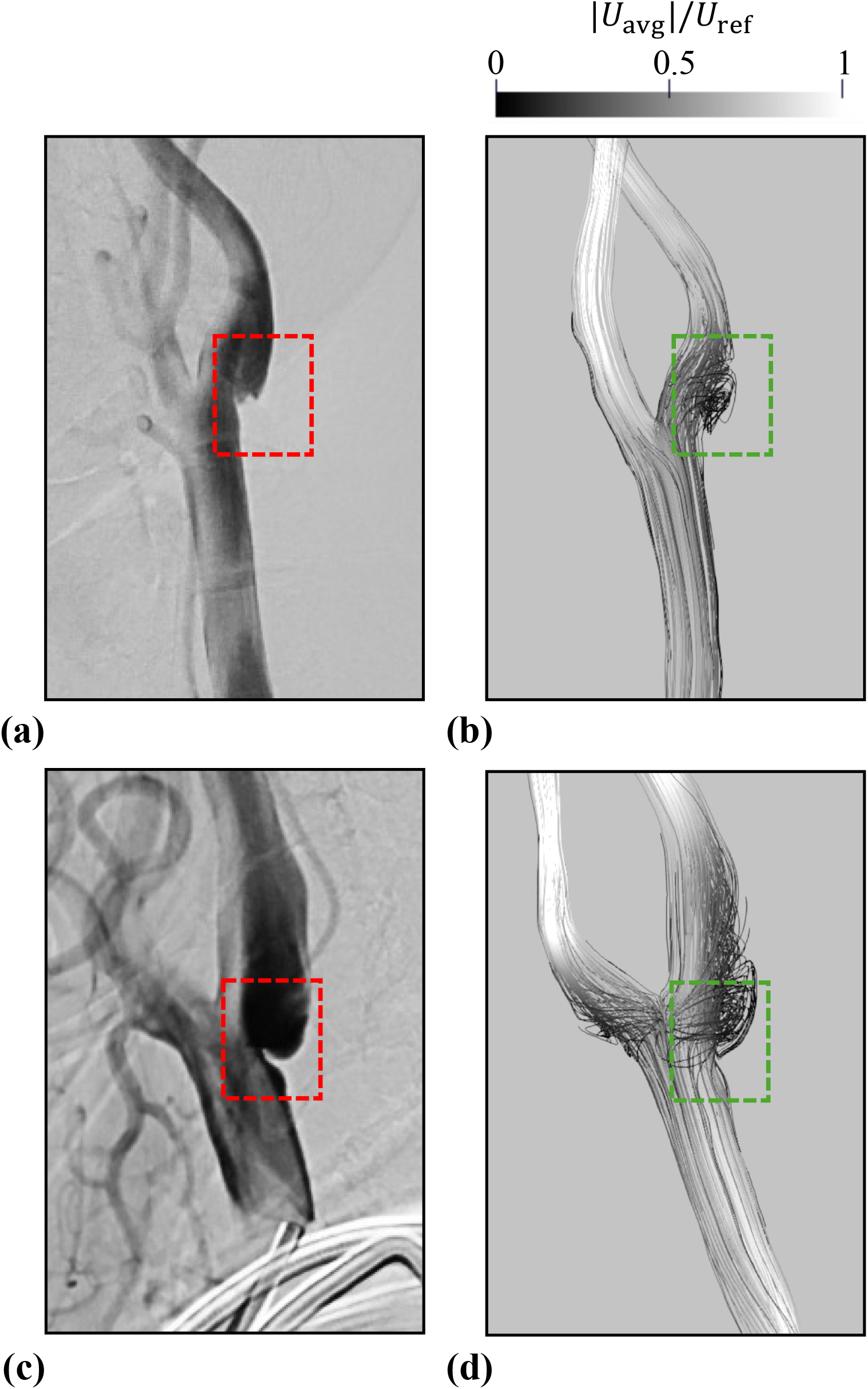
Comparison of angiographic (DSA, frame a and c) and simulated (streamlines, frame b and d) flow patterns for two representative carotid web cases. Streamline color denotes the magnitude of time-averaged velocity normalized by the inlet reference value.

Figure 4 shows representative flow fields and wall-based hemodynamic quantities for a bifurcation with common CaW (web in the bifurcation region), a bifurcation with an atypical CaW (web in ICA), and a normal bifurcation. In the common CaW case, the flow field shows a pronounced recirculation zone distal to the web (black arrow in a1). This downstream disturbance is accompanied by an extended region of low TAWSS, with localized increases in OSI and RRT distributed along the posterior ICA wall. In contrast, the atypical CaW, where the web is within the ICA, shows a different flow pattern. A large recirculation region forms upstream within the carotid bulb (red arrow in a2), while only a small, localized separation is distal to the web (pink arrow in a2). The region proximal to the web shows the lowest TAWSS and the highest OSI and RRT values, reflecting the dominance of the upstream disturbance in this geometry. In the normal bifurcation, only a small flow separation region appears along the outer wall of the carotid bulb (green arrow in a3). Thus, the TAWSS is uniform, OSI minimal, and RRT low across the lumen.

**Figure 4.**
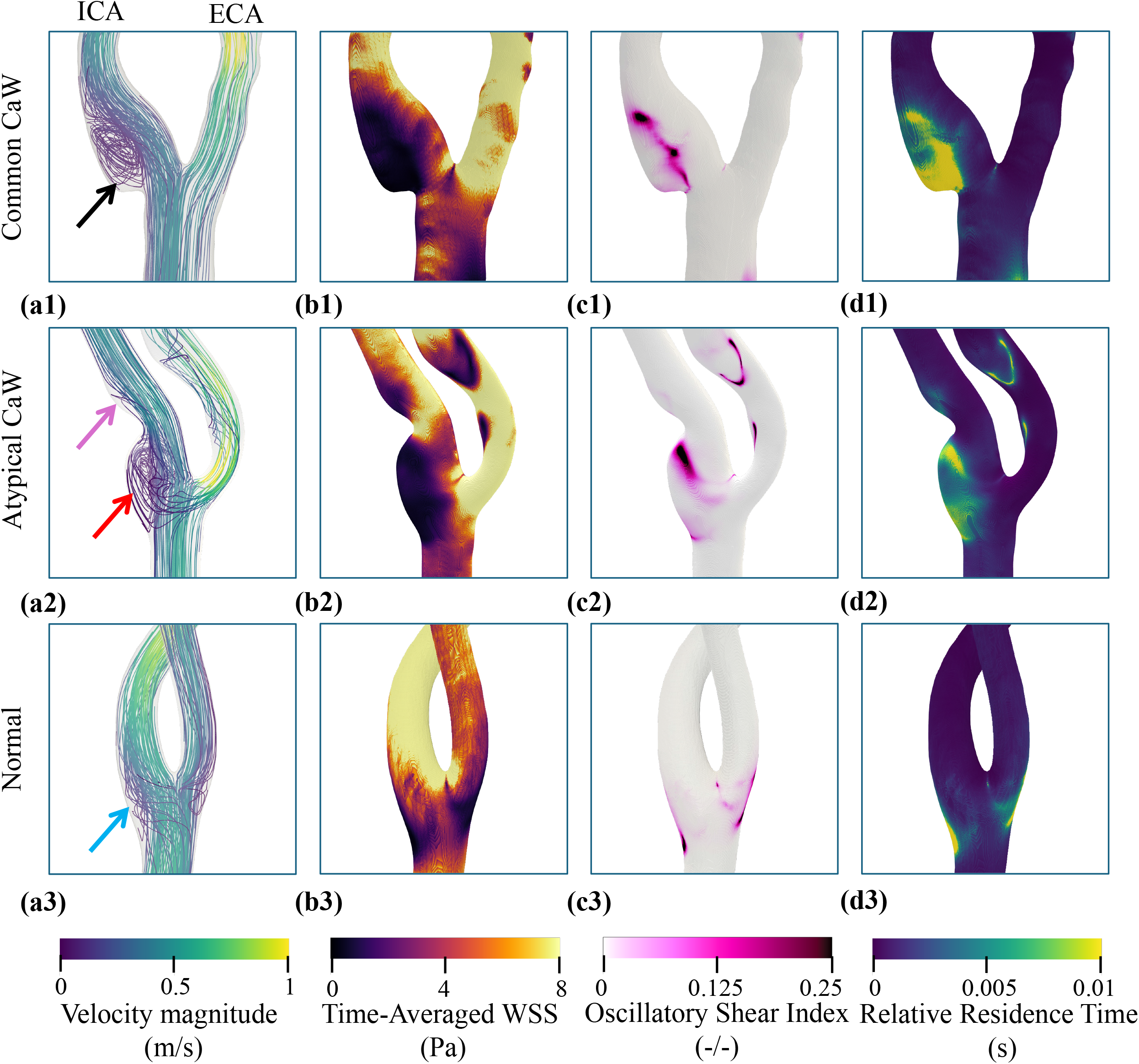
Representative flow fields and wall-based hemodynamic metrics for a common carotid web, an atypical carotid web, and a normal carotid bifurcation. Streamlines are colored by time-averaged velocity magnitude, and wall maps show TAWSS, OSI, and RRT.

### 3.3 Effect of Viscosity Models

We evaluated the sensitivity of the hemodynamic metrics to the choice of Newtonian, Carreau–Yasuda, and Casson viscosity models. The weighted spatial averaged WSS over one cardiac cycle was identical for the three models with peak values differing by less than 2%. The relative timing of systolic and diastolic features is unchanged (see Supplementary Figure S2). The wall-based metrics also showed negligible differences across viscosity models.

### 3.4 Quantitative hemodynamic indices across patient groups

Detailed summary statistics of the weighted hemodynamic metrics and statistical comparisons are provided in Table 2. Overall, the CaW cohort (including both symptomatic and asymptomatic geometries) showed greater inter-patient variability compared with the normal group. For the wall-based hemodynamics metrics, the CaW cohort show a broader distribution, reflecting increased heterogeneity across patient-specific geometries. For example, the TAWSS distribution in the CaW cohort spans a wider range (IQR 2.72 – 8.96 Pa) than in the normal group (IQR 3.09 – 4.56 Pa).

**Table 2.**
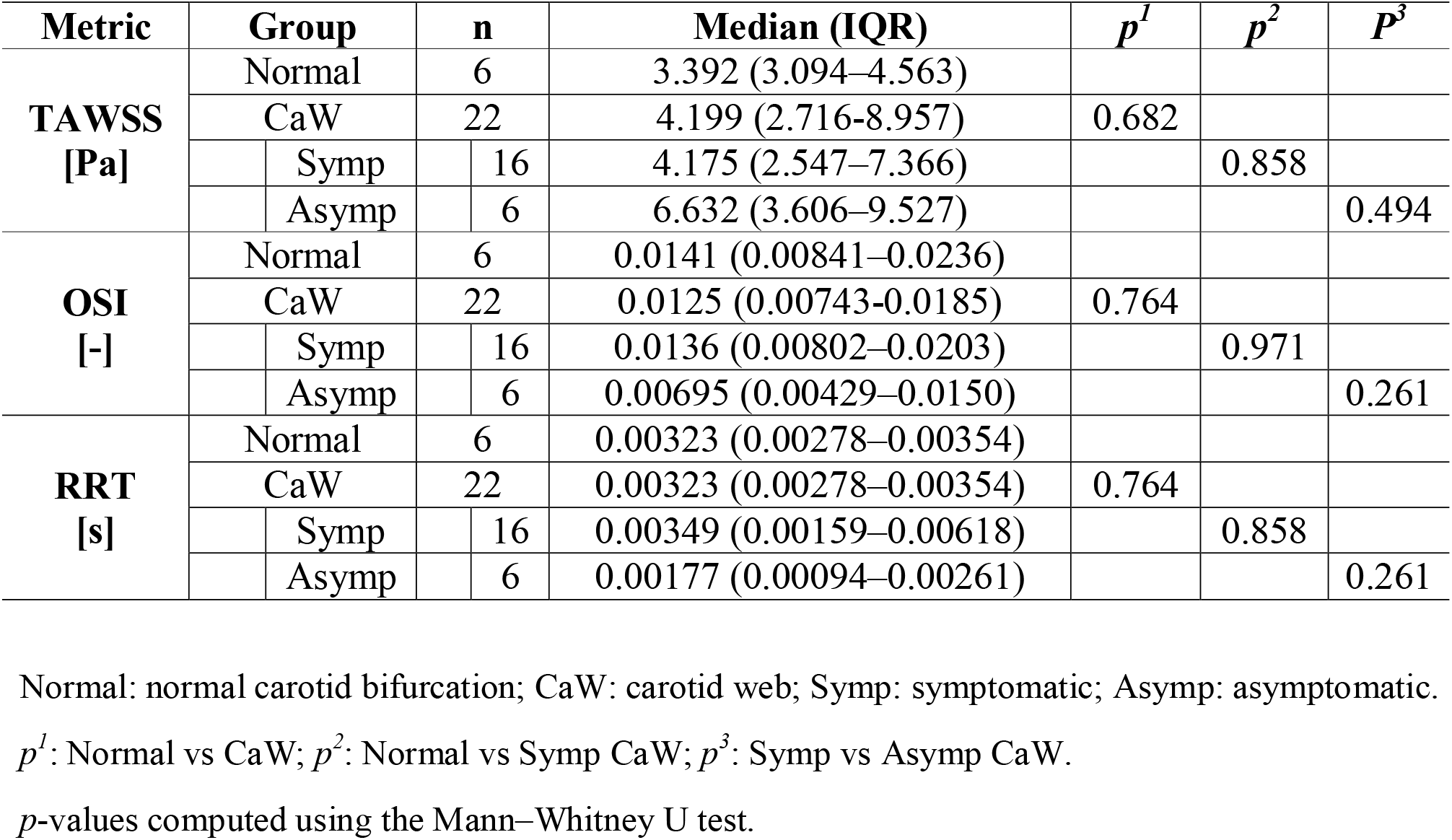
Summary statistics of hemodynamic metrics across patient groups. Values are reported as median and interquartile range (IQR). Pairwise comparisons were performed using the Mann–Whitney U test.

Figure 5 presents the weighted hemodynamic indices for normal carotid bifurcations (Normal), symptomatic carotid webs (Symp CaW), and asymptomatic carotid webs (Asymp CaW). Compared with the normal group, the symptomatic CaW cases show similar median values for the hemodynamic metrics (Symp CaW vs Normal: TAWSS 3.392 Pa vs 4.175 Pa, OSI 0.0141 vs 0.0136, RRT 0.00323 s vs 0.00349 s), and the differences were not statistically significant (TAWSS *p* = 0.858, OSI *p* = 0.971, RRT *p* = 0.858). However, the symptomatic CaW group shows greater variability than the normal group, consistent with the broader variability seen in the entire CaW cohort. Specifically, TAWSS in the symptomatic CaW group spans a wider range (IQR 2.55 – 7.37 Pa) than in the normal group (IQR 3.09 – 4.56 Pa), with several cases extending into the upper range of observed values. Similar increases in spread are also seen for OSI and RRT.

**Figure 5.**
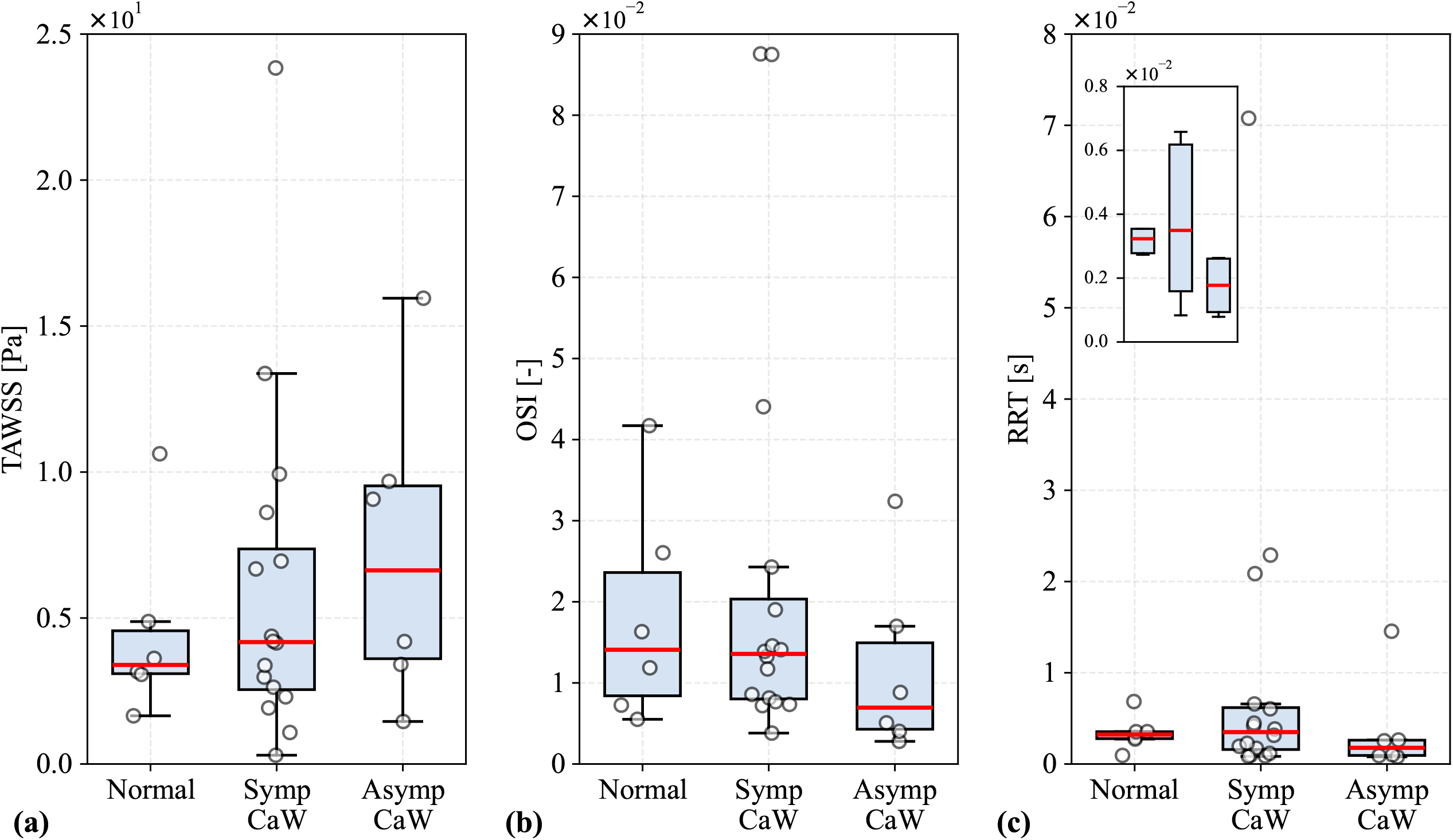
Group-level comparison of weighted hemodynamic indices for normal, carotid web (CaW), and asymptomatic carotid web (Asymp CaW) geometries. Weighted (a) TAWSS, (b) OSI, and (c) RRT are shown for Normal (n = 6), CaW (n = 16), and Asymp CaW (n=6) geometries. Gray boxes indicate interquartile ranges (IQRs), red horizontal lines denote medians, whiskers extend to the most extreme values within 1.5×IQR, and white circles represent individual cases.

A further comparison between symptomatic and asymptomatic CaW geometries shows that symptomatic CaW cases show lower TAWSS and higher OSI and RRT than asymptomatic CaW cases (Symp CaW vs Asymp CaW: TAWSS median 3.392 vs 6.63 Pa, OSI median 0.01441 vs 0.00695, and RRT median 0.00323 vs 0.00177 s), although these differences were not statistically significant (TAWSS *p* = 0.494, OSI *p* = 0.261, RRT *p* = 0.261). Relative to the asymptomatic CaW group, the symptomatic CaW group had an approximately 49% lower TAWSS, 96% higher OSI, and 97% higher RRT, suggesting a trend toward lower shear stress and stronger oscillatory and residence-time characteristics in symptomatic lesions.

## 4. Discussion

Our CFD simulations reproduced the low-velocity recirculation regions distal to the CaWs. Simulated recirculation zone qualitatively matches the regions of delayed contrast clearance in vivo DSA. CaW-induced flow disturbances were distinguishable from those in normal bifurcations. In common CaW geometries (web in the bifurcation region), the shelf-like protrusion generated a pronounced recirculation zone distal to the lesion, which corresponded to a broad region of reduced TAWSS and localized elevations in OSI and RRT. When the web was positioned entirely within the ICA (atypical CaW geometry), the dominant flow disturbance occurred proximally within the bulb rather than downstream. This resulted in low TAWSS and higher OSI and RRT upstream of the lesion with only minimal recirculation distally. Normal carotids differed from the CaW geometries, having consistently low OSI and RRT due to the small physiological flow separation in the bulb. This is consistent with prior studies suggesting that CaWs can promote thrombus formation even in the absence of high-grade stenosis^16,17^.

Newtonian, Carreau–Yasuda, and Casson viscosity models had a negligible effect on the predicted hemodynamic in carotid arteries with CaW. The weighted spatial-averaged WSS waveforms were identical across models, with peak differences below 2%. Similar levels of agreement were seen for TAWSS, OSI and RRT. Moreover, these viscosity variations are only pronounced at low shear rates, i.e., i0.1 s□^1^ and occur in a small portion of the vessel wall (< 3%). These results aligns with prior findings in large artery hemodynamics, including the carotid artery^40–43^, where shear rates are high enough that shear-thinning effects modestly change the viscosity and wall shear stresses. Discrepancies were primarily attributed to differences in viscosity values within low shear regions and did not change these regions nor the recirculation locations. These findings suggest that the choice of blood viscosity model has a limited impact on the hemodynamic conclusions for carotid webs.

Weighted spatial averaged hemodynamic metrics overlap between the CaW and normal bifurcations groups. Moreover, they reduce sensitivity to localized disturbances or intra-luminal transport patterns. Although CaW cases showed greater inter-patient variability, the mean and median values for each metric are like those of the normal group. This lack of separation shows how the bulb expansion and curvature generate regions of low shear and oscillatory flow even in the absence of a web. As a result, the flow disturbances introduced by the CaW may not shift the averaged or wall-integrated hemodynamics from normal carotids in this cohort.

Symptomatic CaW geometries tended to show lower TAWSS and higher OSI and RRT compared with asymptomatic CaWs. These numerical trends suggest a potential association between CaW-related angioarchitecture and altered local hemodynamics. Prior studies have similarly reported that symptomatic CaWs are typically larger and thicker, with increased volume in the region posterior to the web and a more acute angle relative to the carotid wall^44,45^. Differences in CaW morphology (e.g., length and thickness) may influence the extent of flow separation and downstream stasis, thereby contributing to thrombus-prone environments. Collectively, these observations support the concept that CaWs primarily alter downstream flow transport and promote stasis rather than solely affecting near-wall shear magnitude. Consequently, volumetric flow descriptors, particle residence analyses, and Lagrangian transport measures may provide more comprehensive insight into these thrombus-prone conditions. These findings also suggest that incorporating detailed morphologic assessments of CaWs is important to better understand their impact on local hemodynamic parameters and potential thrombotic risk

The limitations of this study are as follows. Firstly, the comparison with angiography was qualitative. DSA provides only projection-based visualization of contrast transport and does not yield quantitative velocity or near-wall shear measurements. Future work incorporating imaging modalities with higher spatial and temporal resolution, or employing contrast-flow quantification techniques, would enable rigorous validation of CaW flow predictions. Secondly, the simulations assumed rigid arterial walls and did not account for vessel compliance or fluid–structure interaction. Although the carotid bifurcation exhibits modest deformation under physiological pressures^46–48^, incorporating wall motion may further refine predictions of near-wall shear and residence characteristics. Thirdly, outflow boundary conditions were prescribed rather than patient-specific, and flow division between the ICA and ECA may vary among individuals. In addition, the present study focused on hemodynamic metrics derived from wall shear stress and did not include a detailed morphological characterization of the web structure (e.g., web length, pocket volume, or protrusion geometry). Future studies combining quantitative morphological assessment with patient-specific CFD simulations may help clarify how variations in CaW angioarchitecture influence local flow patterns and thromboembolic risk. Moreover, although the present cohort is larger than earlier CaW investigations^16–19^, multi-center datasets would enable the assessment of geometric and hemodynamic trends across patient populations. Finally, the normal carotid bifurcations analyzed in this study were from patients that experienced a stroke. Thus, these carotid geometries may have vascular conditions associated with cerebrovascular risk and may partly contribute to the metric overlap between the normal and CaW groups.

## Supporting information

CT protocal

Supplementary figure description

## Data Availability

The data that supports the findings of this study can be obtained from the corresponding author upon reasonable request.

## Author Contribution Statement

XZ: Conceptualization (equal), Methodology (lead), Software (lead), Formal analysis (equal), Investigation (lead), Data curation (lead), Writing - original draft (equal), Writing - review & editing (equal), Project Administration (lead)

FK: Conceptualization (equal), Formal analysis (equal), Writing - original draft (equal), Writing - review & editing (equal), Supervision (equal)

SL: Data curation (lead), Resources (equal), Writing - review & editing (equal)

MRJ: Conceptualization (equal), Writing - original draft (equal), Formal analysis (equal), Writing - review & editing (equal), Funding Acquisition (lead), Supervision (equal)

## Funding

XZ is supported by a Brown University School of Engineering Hibbitt Postdoctoral Research Fellowship and U.S. National Science Foundation (NSF) under Grant No. 2232427. This work used computational resources provided by the Advanced Cyberinfrastructure Coordination Ecosystem: Services & Support (ACCESS) program through allocation PHY250046. The opinions, findings, and conclusions, or recommendations expressed are those of the authors and do not necessarily reflect the views of the funding agencies.

**Supplementary material is available on the journal website**

## References

1. Joux J, Chausson N, Jeannin S, et al. Carotid-Bulb Atypical Fibromuscular Dysplasia in Young Afro-Caribbean Patients With Stroke. Stroke. 2014;45(12):3711–3713. doi:10.1161/STROKEAHA.114.007313

2. Choi PMC, Singh D, Trivedi A, et al. Carotid Webs and Recurrent Ischemic Strokes in the Era of CT Angiography. Am J Neuroradiol. 2015;36(11):2134–2139. doi:10.3174/ajnr.A4431

3. Chen H, Colasurdo M, Costa M, Nossek E, Kan P. Carotid webs: a review of pathophysiology, diagnostic findings, and treatment options. J NeuroInterventional Surg. 2024;16(12):1294–1299. doi:10.1136/jnis-2023-021243

4. Haussen DC, Grossberg JA, Bouslama M, et al. Carotid Web (Intimal Fibromuscular Dysplasia) Has High Stroke Recurrence Risk and Is Amenable to Stenting. Stroke. 2017;48(11):3134–3137. doi:10.1161/STROKEAHA.117.019020

5. Sajedi PI, Gonzalez JN, Cronin CA, et al. Carotid Bulb Webs as a Cause of “Cryptogenic” Ischemic Stroke. Am J Neuroradiol. 2017;38(7):1399–1404. doi:10.3174/ajnr.A5208

6. Zhang AJ, Dhruv P, Choi P, et al. A Systematic Literature Review of Patients With Carotid Web and Acute Ischemic Stroke. Stroke. 2018;49(12):2872–2876. doi:10.1161/STROKEAHA.118.021907

7. Olindo S, Chausson N, Signate A, et al. Stroke Recurrence in First-Ever Symptomatic Carotid Web: A Cohort Study. J Stroke. 2021;23(2):253–262. doi:10.5853/jos.2020.05225

8. Khan F, Kala N, Chang K, et al. In-hospital recurrent stroke in ipsilateral carotid web patients undergoing thrombectomy. Ann Clin Transl Neurol. 2024;11(9):2450–2456. doi:10.1002/acn3.52161

9. Guglielmi V, Compagne KCJ, Sarrami AH, et al. Assessment of Recurrent Stroke Risk in Patients With a Carotid Web. JAMA Neurol. 2021;78(7):826–833. doi:10.1001/jamaneurol.2021.1101

10. Park CC, Sayed RE, Risk BB, et al. Carotid webs produce greater hemodynamic disturbances than atherosclerotic disease: a DSA time–density curve study. J NeuroInterventional Surg. 2022;14(7):729–733. doi:10.1136/neurintsurg-2021-017588

11. Damiani Monteiro M, Tarek MA, Martins PN, et al. Carotid web catheter angiography hemodynamic parameters. J NeuroInterventional Surg. 2025;17(8):843–847. doi:10.1136/jnis-2024-021948

12. El Sayed R, Park CC, Shah Z, et al. Assessment of Complex Flow Patterns in Patients With Carotid Webs, Patients With Carotid Atherosclerosis, and Healthy Subjects Using 4D Flow MRI. J Magn Reson Imaging. 2024;59(6):2001–2010. doi:10.1002/jmri.29013

13. Bae T, Ko JH, Chung J. Turbulence Intensity as an Indicator for Ischemic Stroke in the Carotid Web. World Neurosurg. 2021;154:e443–e457. doi:10.1016/j.wneu.2021.07.049

14. Liu X, Song P, Gao Q, Dai M, Rao J, Wen J. Impact on hemodynamics in carotid arteries with carotid webs at different locations: A Numerical Study Integrating Thrombus Growth Model. Comput Methods Programs Biomed. 2024;243:107926. doi:10.1016/j.cmpb.2023.107926

15. Moghadasi K, Ghayesh MH, Hu E, Li J. Nonlinear biomechanics of diseased carotid arteries. Int J Eng Sci. 2024;199:104070. doi:10.1016/j.ijengsci.2024.104070

16. Compagne KCJ, Dilba K, Postema EJ, et al. Flow Patterns in Carotid Webs: A Patient-Based Computational Fluid Dynamics Study. AJNR Am J Neuroradiol. 2019;40(4):703–708. doi:10.3174/ajnr.A6012

17. El Sayed R, Lucas CJ, Cebull HL, et al. Subjects with carotid webs demonstrate pro-thrombotic hemodynamics compared to subjects with carotid atherosclerosis. Sci Rep. 2024;14(1):10092. doi:10.1038/s41598-024-60666-7

18. Ren S, Liu Q, Chen Z, Deng X, Sun A, Luan J. Hemodynamic evaluation of endarterectomy and stenting treatments for carotid web. Front Cardiovasc Med. 2022;9. doi:10.3389/fcvm.2022.993037

19. Ozaki D, Endo T, Suzuki H, et al. Carotid web leads to new thrombus formation: computational fluid dynamic analysis coupled with histological evidence. Acta Neurochir (Wien). 2020;162(10):2583–2588. doi:10.1007/s00701-020-04272-2

20. Yilmaz F, Gundogdu M. A critical review on blood flow in large arteries; relevance to blood rheology, viscosity models, and physiologic conditions. Korea-Aust Rheol J. 2008;20:197–211.

21. Alexy T, Detterich J, Connes P, et al. Physical Properties of Blood and their Relationship to Clinical Conditions. Front Physiol. 2022;13. doi:10.3389/fphys.2022.906768

22. Beris AN, Horner JS, Jariwala S, Armstrong MJ, Wagner NJ. Recent advances in blood rheology: a review. Soft Matter. 2021;17(47):10591–10613. doi:10.1039/D1SM01212F

23. Liu S, Wang S, Tian H, et al. Comparison of blood viscosity models in different degrees of carotid artery stenosis. PeerJ. 2025;13:e19336. doi:10.7717/peerj.19336

24. Spatial comparison between wall shear stress measures and porcine arterial endothelial permeability. doi:10.1152/ajpheart.00897.2003

25. Malota Z, Glowacki J, Sadowski W, Kostur M. Numerical analysis of the impact of flow rate, heart rate, vessel geometry, and degree of stenosis on coronary hemodynamic indices. BMC Cardiovasc Disord. 2018;18(1):132. doi:10.1186/s12872-018-0865-6

26. Song J, Gao S, Xie E, et al. Systematic Review of the Application of Computational Fluid Dynamics for Adult Aortic Diseases. Rev Cardiovasc Med. 2023;24(12):355. doi:10.31083/j.rcm2412355

27. Lopes D, Puga H, Teixeira J, Lima R. Blood flow simulations in patient-specific geometries of the carotid artery: A systematic review. J Biomech. 2020;111:110019. doi:10.1016/j.jbiomech.2020.110019

28. Azar D, Torres WM, Davis LA, et al. Geometric determinants of local hemodynamics in severe carotid artery stenosis. Comput Biol Med. 2019;114:103436. doi:10.1016/j.compbiomed.2019.103436

29. Carallo C, Lucca LF, Ciamei M, Tucci S, de Franceschi MS. Wall shear stress is lower in the carotid artery responsible for a unilateral ischemic stroke. Atherosclerosis. 2006;185(1):108–113. doi:10.1016/j.atherosclerosis.2005.05.019

30. Malek AM, Alper SL, Izumo S. Hemodynamic Shear Stress and Its Role in Atherosclerosis. JAMA. 1999;282(21):2035–2042. doi:10.1001/jama.282.21.2035

31. Ku DN, Giddens DP, Zarins CK, Glagov S. Pulsatile flow and atherosclerosis in the human carotid bifurcation. Positive correlation between plaque location and low oscillating shear stress. Arterioscler Off J Am Heart Assoc Inc. 1985;5(3):293–302. doi:10.1161/01.ATV.5.3.293

32. Jackson ML, Bond AR, George SJ. Mechanobiology of the endothelium in vascular health and disease: in vitro shear stress models. Cardiovasc Drugs Ther. 2023;37(5):997–1010. doi:10.1007/s10557-022-07385-1

33. Lee SW, Antiga L, Spence JD, Steinman DA. Geometry of the Carotid Bifurcation Predicts Its Exposure to Disturbed Flow. Stroke. 2008;39(8):2341–2347. doi:10.1161/STROKEAHA.107.510644

34. Yamamoto H, Yabuta T, Negi Y, et al. Measurement of human blood viscosity a using Falling Needle Rheometer and the correlation to the Modified Herschel-Bulkley model equation. Heliyon. 2020;6(9):e04792. doi:10.1016/j.heliyon.2020.e04792

35. Horner JS. An Experimental and Theoretical Investigation of Blood Rheology. Ph.D. University of Delaware; 2020. Accessed March 24, 2025. https://www.proquest.com/docview/2461026320/abstract/719DFCF11E60486DPQ/1

36. Furukawa K, Abumiya T, Sakai K, et al. Measurement of human blood viscosity by an electromagnetic spinning sphere viscometer. J Med Eng Technol. 2016;40(6):285–292. doi:10.1080/03091902.2016.1181216

37. Kim BJ, Lee SY, Jee S, Atajanov A, Yang S. Micro-Viscometer for Measuring Shear-Varying Blood Viscosity over a Wide-Ranging Shear Rate. Sensors. 2017;17(6):6. doi:10.3390/s17061442

38. Skalak R, Keller SR, Secomb TW. ASME Centennial Historical Perspective Paper: Mechanics of Blood Flow. J Biomech Eng. 1981;103(2):102–115. doi:10.1115/1.3138253

39. Ferguson GG, Eliasziw M, Barr HWK, et al. The North American Symptomatic Carotid Endarterectomy Trial. Stroke. 1999;30(9):1751–1758. doi:10.1161/01.STR.30.9.1751

40. Gharahi H, Zambrano BA, Zhu DC, DeMarco JK, Baek S. Computational fluid dynamic simulation of human carotid artery bifurcation based on anatomy and volumetric blood flow rate measured with magnetic resonance imaging. Int J Adv Eng Sci Appl Math. 2016;8(1):40. doi:10.1007/s12572-016-0161-6

41. Morbiducci U, Gallo D, Massai D, et al. On the importance of blood rheology for bulk flow in hemodynamic models of the carotid bifurcation. J Biomech. 2011;44(13):2427–2438. doi:10.1016/j.jbiomech.2011.06.028

42. Lee SW, Steinman DA. On the relative importance of rheology for image-based CFD models of the carotid bifurcation. J Biomech Eng. 2007;129(2):273–278. doi:10.1115/1.2540836

43. A study of wall shear stress in 12 aneurysms with respect to different viscosity models and flow conditions. J Biomech. 2013;46(16):2802–2808. doi:10.1016/j.jbiomech.2013.09.004

44. Tabibian BE, Parr M, Salehani A, et al. Morphological characteristics of symptomatic and asymptomatic carotid webs. J Neurosurg. 2022;137(6):1727–1732. doi:10.3171/2022.2.JNS212310

45. Bala F, Alhabli I, Singh N, et al. Relationship between carotid web morphology on CT angiography and stroke: A pooled multicenter analysis. Int J Stroke. 2024;19(9):1046–1052. doi:10.1177/17474930241264141

46. Albadawi M, Abuouf Y, Elsagheer S, Sekiguchi H, Ookawara S, Ahmed M. Influence of Rigid–Elastic Artery Wall of Carotid and Coronary Stenosis on Hemodynamics. Bioengineering. 2022;9(11):708. doi:10.3390/bioengineering9110708

47. Dong J, Sun Z, Inthavong K, Tu J. Fluid-structure interaction analysis of the left coronary artery with variable angulation. Comput Methods Biomech Biomed Engin. 2015;18(14):1500–1508. doi:10.1080/10255842.2014.921682

48. Bantwal A, Singh A, Menon AR, Kumar N. Pathogenesis of atherosclerosis and its influence on local hemodynamics: A comparative FSI study in healthy and mildly stenosed carotid arteries. Int J Eng Sci. 2021;167:103525. doi:10.1016/j.ijengsci.2021.103525

