## Supplementary material for "A Patient-Specific CFD Study of Carotid Webs: Hemodynamic Analysis and the Role of Blood Viscosity": CT protocal

### BUH –BRAIN AND CAROTID CTA

**Indications: carotid/cerebral artery stenosis or blockage; non-trauma**

IV placement/management included in the protocol

| | |
|---|---|
| **Position/Landmark** | Supine head first or feet first |
| **Topogram Direction** | Craniocaudal |
| **Scan Type** | Helical |
| **First Helical Set**<br>Slice Thickness/ Spacing<br>Algorithm<br>Recon Destination | recon / body part / thickness/spacing / algorithm / recon destination<br>**non angled head** / 5mm x 5mm / standard / pacs<br>thin brain / .6mm x .6mm / standard / mpr/pacs/RapidAI |
| **Second Helical Set**<br>Slice Thickness/ Spacing<br>Algorithm<br>Recon Destination | recon / body part / thickness/spacing / algorithm / recon destination<br>thin cta carotid/brain / .6mm x .6mm / soft / mpr/pacs<br>**axial neck brain cta** / 1.2mm x 1.2mm / standard / pacs/RapidAI |
| **Scan Start / End Locations**<br><br>**DFOV** | brain: 1cm inferior to skull base/skull vertex; 25cm<br>cta neck brain: 1cm inferior to aortic arch/skull vertex; 25cm |
| **IV Contrast Volume / Type / Rate** | 80mL Iohexol (Omnipaque 350) / 4mL per second |
| **Scan Delay** | Smart Prep at Aortic Arch<br>HU threshold is +100 |
| **2D/3D Technique Used** | **Non Con**: 5mm x 5mm **axial, coronal, and sagittal brain reformats** in respect to the glabello-meatal plane, average mode<br><br>**CTA**: **axial reformats** 10mm x 3mm, mip mode<br>**coronal reformats** 1mm x 1mm, mip mode<br>**sagittal reformats** 1mm x 1mm, mip mode |
| **Comments:** | |
| **Images required in PACS and Archive** | Scouts, 5mm x 5mm head, 5mm x 5mm axial nc brain, 5mm x 5mm coronal and sagittal nc brain, 1.2mm x 1.2mm axial neck and brain cta, 10mm x 3mm axial neck brain cta mip, 1mm x 1mm sagittal neck brain cta mip, 1mm x 1mm coronal neck brain cta mip, thin data, Dose Report |

reviewed October 1, 2025 sc
