## Supplementary figures and images for "A Patient-Specific CFD Study of Carotid Webs: Hemodynamic Analysis and the Role of Blood Viscosity"

### Supplementary Figure 1

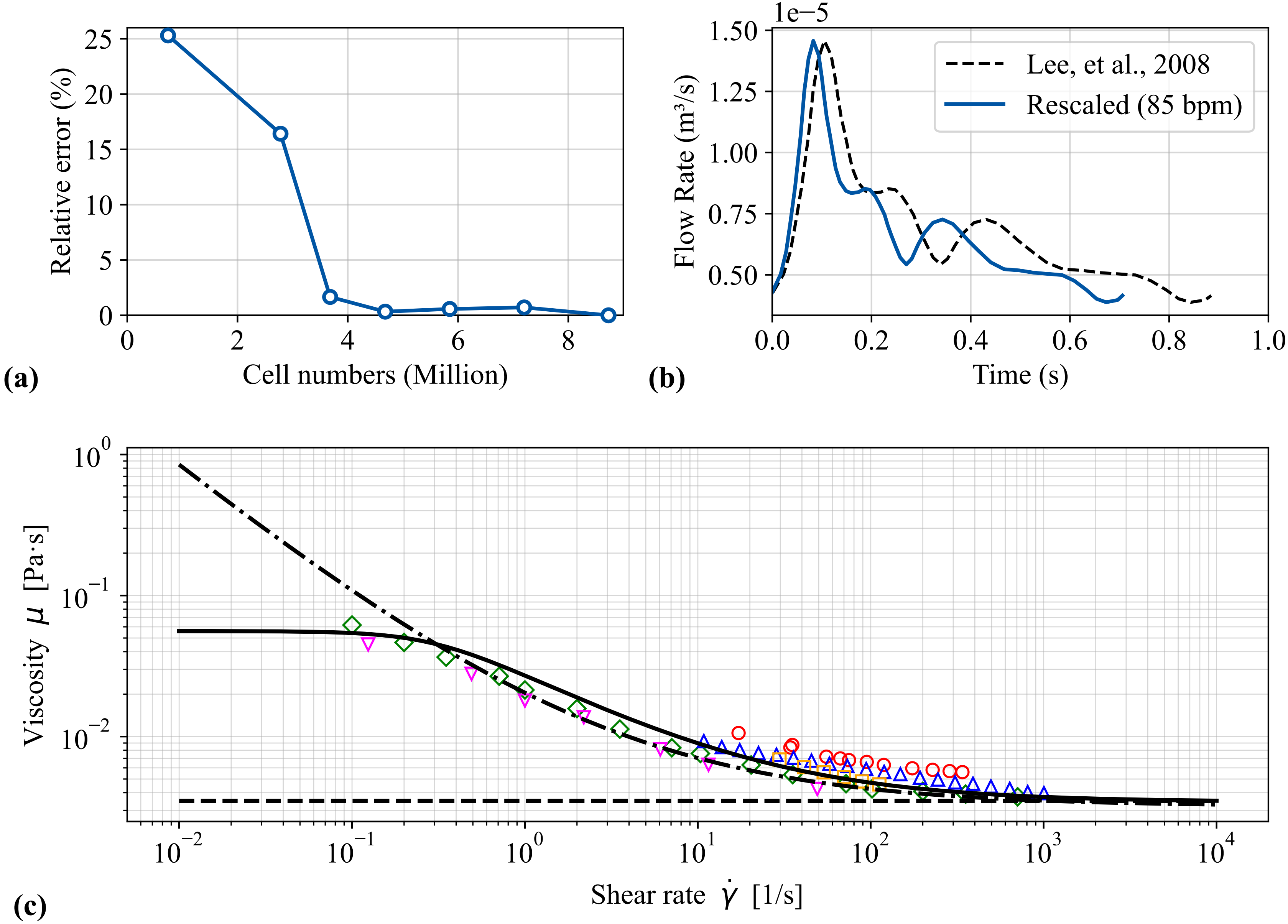

### Supplementary Figure 2

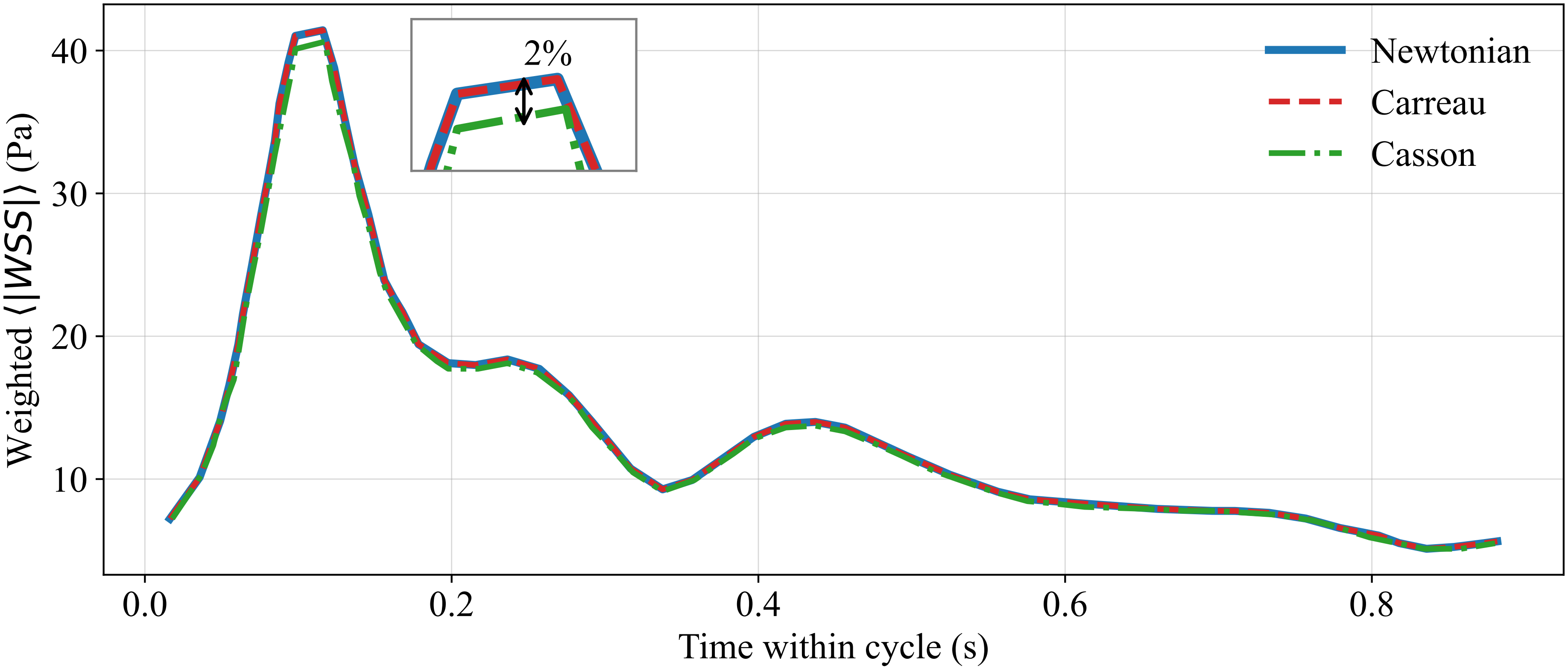
