## Supplementary figure description for "A Patient-Specific CFD Study of Carotid Webs: Hemodynamic Analysis and the Role of Blood Viscosity"

**Supplementary figures**

Figure S1. (a) Mesh resolution study illustrating the relative error of the time- and spatially averaged wall shear stress as a function of total cell number. Convergence is seen beyond approximately 4 million computational elements. (b) Prescribed pulsatile inlet volumetric flow waveform used as the inflow boundary condition in the simulations. The dashed line shows the waveform reported in published physiological measurements, and the solid line shows the waveform rescaled to the corresponding heart rate, with an example shown for 85 bpm. (c) Blood viscosity as a function of shear rate. Viscosity predicted by the Newtonian (black dashed line), Carreau–Yasuda (black solid line), and Casson (black dash-dot line) models compared with experimental data from multiple studies (red circles: Yamamoto, 2020^34^; green diamonds: Horner, 2020^35^: orange squares: Furukawa, 2016^36^; blue upward triangles: Kim, 2017^37^; and magenta downward triangles: Skalak, 1981^38^). As expected, the Newtonian model (dashed line) shows constant viscosity and does not capture shear-thinning behavior. Both Carreau–Yasuda (solid black line) and Casson (dash-dot black line) models reproduce the characteristic decrease in apparent viscosity with increasing shear rate, closely following the experimental trend between 10-1000 s^-1^. At shear rates < 0.1 s^-1^, the Casson model predicts higher viscosities due to the influence of yield stress, while the Carreau–Yasuda model shows a smoother transition between the low- and high-shear plateaus.

Figure S2. Weighted wall shear stress (WSS) over one cardiac cycle for three viscosity models for a representative carotid web case. The Newtonian, Carreau–Yasuda, and Casson formulations produce nearly identical WSS waveforms, with peak differences below 2%.
